# Visuospatial neglect: Recovery & Functional outcome after 6 months

**DOI:** 10.1101/2021.03.29.21254555

**Authors:** Margaret Jane Moore, Kathleen Vancleef, M. Jane Riddoch, Celine R Gillebert, Nele Demeyere

## Abstract

**Background/Objective:** This study aims to investigate how complex visuospatial neglect behavioural phenotypes predict long-term outcomes, both in terms of neglect recovery and broader functional outcomes.

**Methods:** This study presents a secondary cohort study of acute and 6 month follow up data from 400 stroke survivors who completed the Oxford Cognitive Screen’s Cancellation Task. At follow-up, patients also completed the Stroke Impact Scale questionnaire. These data were analysed to identify whether any specific combination of neglect symptoms is more likely to result in long-lasting neglect or higher levels of functional impairment, therefore warranting more targeted rehabilitation.

**Results:** Overall, 98/142(69%) neglect cases recovered by follow-up and there was no significant difference in the persistence of egocentric/allocentric (X^2^(1)=0.66, *p=0.418*) or left/right neglect (X^2^(2)=0.781, *p= 0.677)*. Egocentric neglect was found to follow a proportional recovery pattern with all patients demonstrating a similar level of improvement over time. Conversely, allocentric neglect followed a non-proportional recovery pattern with chronic neglect patients exhibiting a slower rate of improvement than those who recovered. A multiple regression analysis revealed that the initial severity of acute allocentric, but not egocentric, neglect impairment acted as a significant predictor of poor long-term functional outcomes (F(9,383)=3.96, *p<0.001*, R^2^=0.066).

**Conclusions:** Our findings call for systematic neuropsychological assessment of both egocentric and allocentric neglect following stroke, as the occurrence and severity of these conditions may help predict recovery outcomes.

## Introduction

Visuospatial neglect is a common neuropsychological syndrome characterised by consistently lateralised perceptual deficits ^1,2^. The neglect syndrome is represented by a highly heterogeneous group of symptoms and contains many subtypes ^3–5^. Although visuospatial neglect is a common consequence of stroke, it is not yet clear whether different subtypes of neglect follow similar recovery trajectories or whether neglect subtypes are differentially associated with poor long-term functional outcomes.

The occurrence of post-stroke cognitive impairments has been strongly associated with reduced quality of life throughout recovery^6–9^. However, not all cognitive deficits appear to affect quality of life and functional recovery to the same extent. Previous research has demonstrated that patients who experience visuospatial neglect following stroke are significantly more likely to report higher levels of functional impairment and lower quality of life than patients without visuospatial neglect impairment ^6,7,10^. This particularly robust effect has been documented across multiple different timepoints using a wide range of functional outcome measures. For example, Jehkonen et al.^6^ found that performance on the Behavioural Inattention Test was the single best predictor (compared to hemianopia, age, and verbal memory) of poor functional outcome at 3, 6 and 12 month follow-up appointments. Cherney et al.^10^ determined that higher neglect severity was predictive of lower Functional Independence Measure scores at admission, discharge, and 3 month follow-up. Katz et al.^7^ found that acute neglect was associated with lower scores on various activities of daily living measurements as well as on standardised cognitive measures throughout the first five months following stroke. Similarly, Buxbaum et al.^11^ concluded that the occurrence of neglect deficits predicted poor recovery outcome following stroke over and above general stroke severity metrics. These findings demonstrate that the occurrence of visuospatial neglect acts as a significant predictor of functional recovery outcome.

However, neglect is not a unitary syndrome. Following stroke, patients can exhibit visuospatial neglect within a self-centred (egocentric) and/or object-centred reference frame (allocentric neglect)^3,3,4,12–16^. For example, a patient with egocentric neglect might fail to notice objects presented on their neglected side while a patient with allocentric neglect might fail to perceive features appearing on the neglected side of individual objects, regardless of where these objects are presented in space^17^. While egocentric and allocentric neglect do frequently co-occur, these conditions have been demonstrated to represent doubly-dissociated, independent cognitive impairments^3,14,18,19^. Importantly, additional subtypes of neglect have been documented. Patients can selectively exhibit neglect within peri-personal (near space) and extra-personal space ^20–23^. Patients have also been found to exhibit spatial attentional biases in additional sensory modalities including auditory neglect^24–26^ and motor neglect ^27–29^. However, standardised neuropsychological tests which can reliably detect and differentiate between these additional neglect subtypes are not commonly employed in clinical environments^30–32^. For this reason, this analysis will focus on exploring the recovery trajectories of egocentric and allocentric neglect. The present study aims to investigate only differences between egocentric and allocentric neglect with the implication being that if these two subtypes recover differentially, additional research will be needed to investigate whether these differences are also present within other subtypes of neglect.

Previous research has suggested that patients with egocentric and allocentric neglect may exhibit differing levels of functional impairment. Bickerton et al.^3^ found that patients with allocentric neglect scored significantly lower (e.g. worse functional performance) on the Barthel Index activities of daily life measurement^33^ compared to patients with egocentric neglect at a single, subacute time point. In this study, patients with both allocentric and egocentric neglect also reported significantly higher levels of depression on the Hospital Anxiety and Depression Scale than patients with either egocentric or allocentric neglect alone ^3^.

Similarly, although neglect is most commonly thought of as a left-lateralised impairment occurring following unilateral right-hemisphere damage, recent research has demonstrated that right-lateralised neglect impairments also frequently occur following stroke ^34–39^. Patients with left and right neglect may exhibit differing levels of functional impairment. Ten Brink et al.^40^ investigated the relationship between neglect lateralisation and performance on various cognitive and physical independence measures in a cohort of 335 acute stroke survivors. This study found that left-lateralised neglect impairment was more severe than right-lateralised neglect as assessed by both neuropsychological and observational measures. However, patients with right neglect exhibited lower scores on the Mini Mental State Examination^41,42^ than patients without neglect (potentially through a co-occurring language deficit) and were more likely to exhibit impaired balance than patients with left neglect ^40^. These findings suggest that the association between neglect and functional impairment may depend on the reference frame and lateralisation of neglect. However, these studies only employed data from a single timepoint acutely post stroke.

Overall, previous studies which have tracked neglect recovery over time have found that the majority of neglect cases recover within the first six months following stroke ^36,39,43^. Nijboer et al.^43^ found that time post-stroke was a key and independent predictor of visuospatial neglect recovery, with 54% of patients recovering within the first 12 weeks and around 60% recovered within the first year. A subsequent study found that clinical characteristics including neglect severity, stroke severity, and severity of comorbid stoke deficits did not act as significant predictors of neglect recovery over the first 26 weeks following stroke^44^. These studies provide important insight into the recovery trajectory of neglect as a whole, but do not distinguish between egocentric and allocentric neglect deficits. Several previous studies have investigated how different neglect subtypes recover over time. Stone et al^39^ in a group of 68 patients with neglect, found that neglect following right hemisphere damage (N=34) was significantly slower to improve and less likely to fully recover than neglect following left hemisphere lesions (N=34) over the first six months following stroke. Demeyere & Gillebert^36^ found that of the 160 patients with follow up data 81% of egocentric neglect (11 impaired at follow up versus 55 impaired at the stage) and 71% of allocentric neglect cases (10 chronic versus 39 acute) recovered within six months following stroke. Overall, a high proportion of visuospatial neglect cases were found to spontaneously recover within the first six months following stroke^14,39^, but this relationship may not be equivalently true for different neglect lateralisations and reference frames.

Previous research has suggested that some stroke-related deficits follow a proportional recovery rule in which the amount of improved function is proportional to the initial degree of function loss^45,46^. However, it is not yet clear whether egocentric and allocentric visuospatial neglect follow this proportional recovery pattern^47,48^. If neglect impairments follow a proportional recovery rule, neglect cases which are initially more severe should be less likely to fully recover than less severe cases^45,46^. Conversely, if neglect does not follow a proportional recovery pattern, the initial severity of neglect would not be expected to differentiate between patients who do and do not recover over time. Similarly, it is important to investigate differences in rates of improvement over time across different neglect subtypes. For example, it is possible that patients with milder deficits improve gradually over time while patients with severe impairments do not experience any improvement. Alternatively, it is possible that all patients with neglect exhibit the same rate of change in severity over time, regardless of the severity of their initial deficits. It is therefore critically important to better characterise recovery trajectories in different neglect subtypes in order to identify the specific patients who are least likely to fully recover.

Previous investigations into neglect recovery trajectories have not considered the interaction between neglect subtype and lateralisation when attempting to elucidate the predictive relationships between these acute factors and long-term recovery outcomes. Here, for the first time, we present data from a large and representative longitudinal sample of stroke survivors to attempt to disentangle how complex acute neglect behavioural phenotypes predict longer-term recovery outcomes, both in terms of long-lasting neglect symptoms as well as broader functional outcomes related to activities of daily life and participation. The present study aims to investigate which specific attributes of acute neglect impairment are related to a low likelihood of neglect recovery 6 months later and which are associated with worse functional outcomes. This in turn, may help identify which specific patients are the most likely to benefit from targeted rehabilitation strategies.

## Materials and Methods

### Participants

This project represents a secondary analysis of data collected within the Oxford Cognitive Screen (OCS)^49^, OCS-Tablet, and OCS-Care^50^ studies from 2015-2019. These study protocols were reviewed and approved by the National Research Ethics Committee (UK) (References: 11/WM/0299, 14/LO/0648, and 12/WM/00335 respectively). Each study included acute cognitive screening (T1) and 6 month follow-ups (T2). Patients were included in this analysis if Cancellation Task data was available from both timepoints and they had completed the Stroke Impact Scale (SIS) at timepoint 2 (Figure 1). A total of 400 patients (mean age=70.6(SD= 12.2), 47% female, mean years of education =12 years(SD= 2.78), 6% left handed) were included in this study. As reported by patient clinical notes, this sample included 322 ischemic, 57 haemorrhagic, and 21 not reported stroke types (Figure 1). Lesion sides were reported as 197 right, 157 left, 29 bilateral, and 17 not reported. The average stroke-test interval was 9.9(SD=11.9) days at T1. Table 1 presents a full summary of demographic and stroke information for the patients in this study with and without significant neglect impairment.

**Figure 1:**
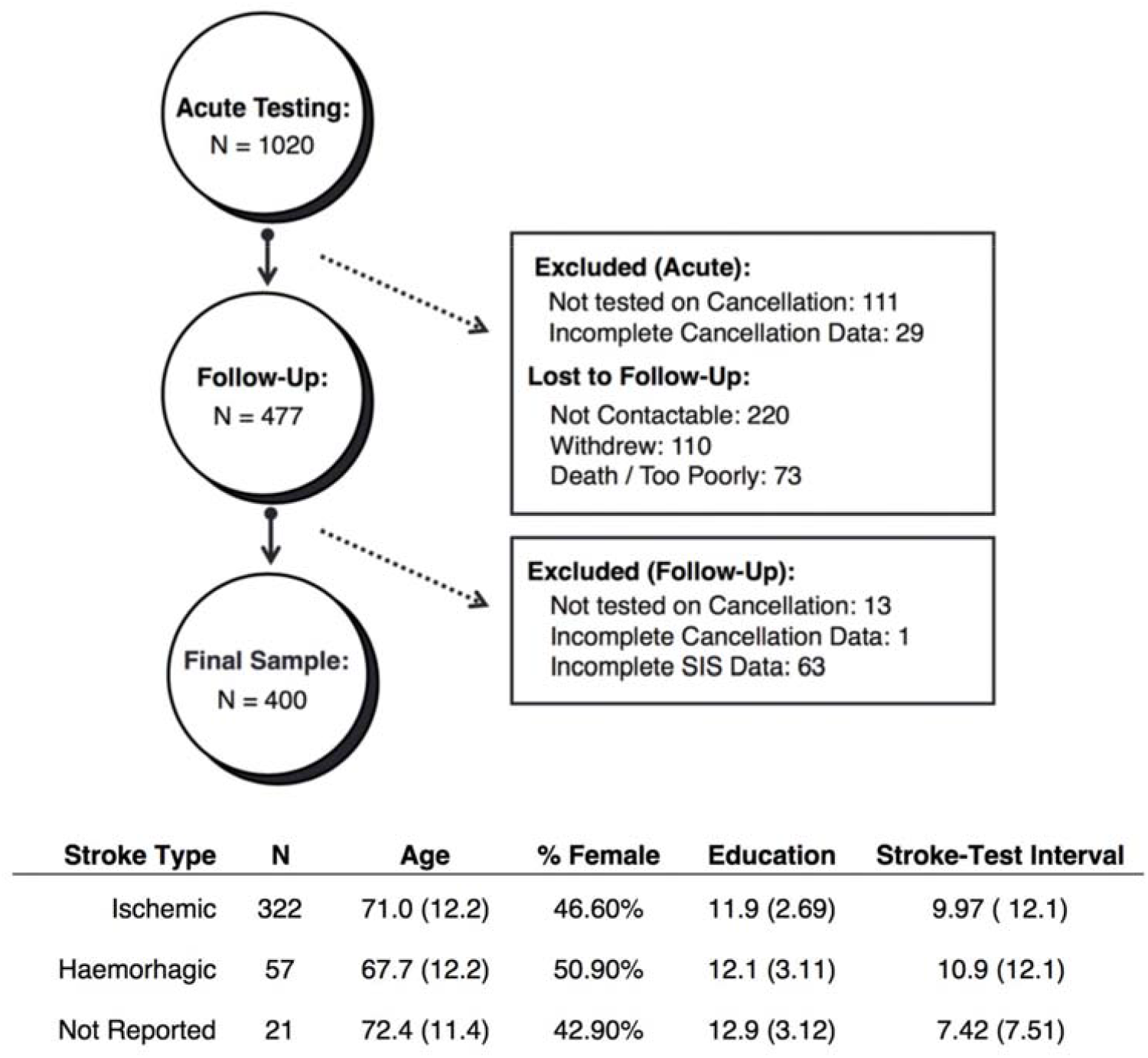
Inclusion/Exclusion Criteria. A summary of patient exclusion criteria and dropout counts at each stage along with demographic details for the final sample (n = 400). Means are reported followed by standard deviations in parentheses.

**Table One:**
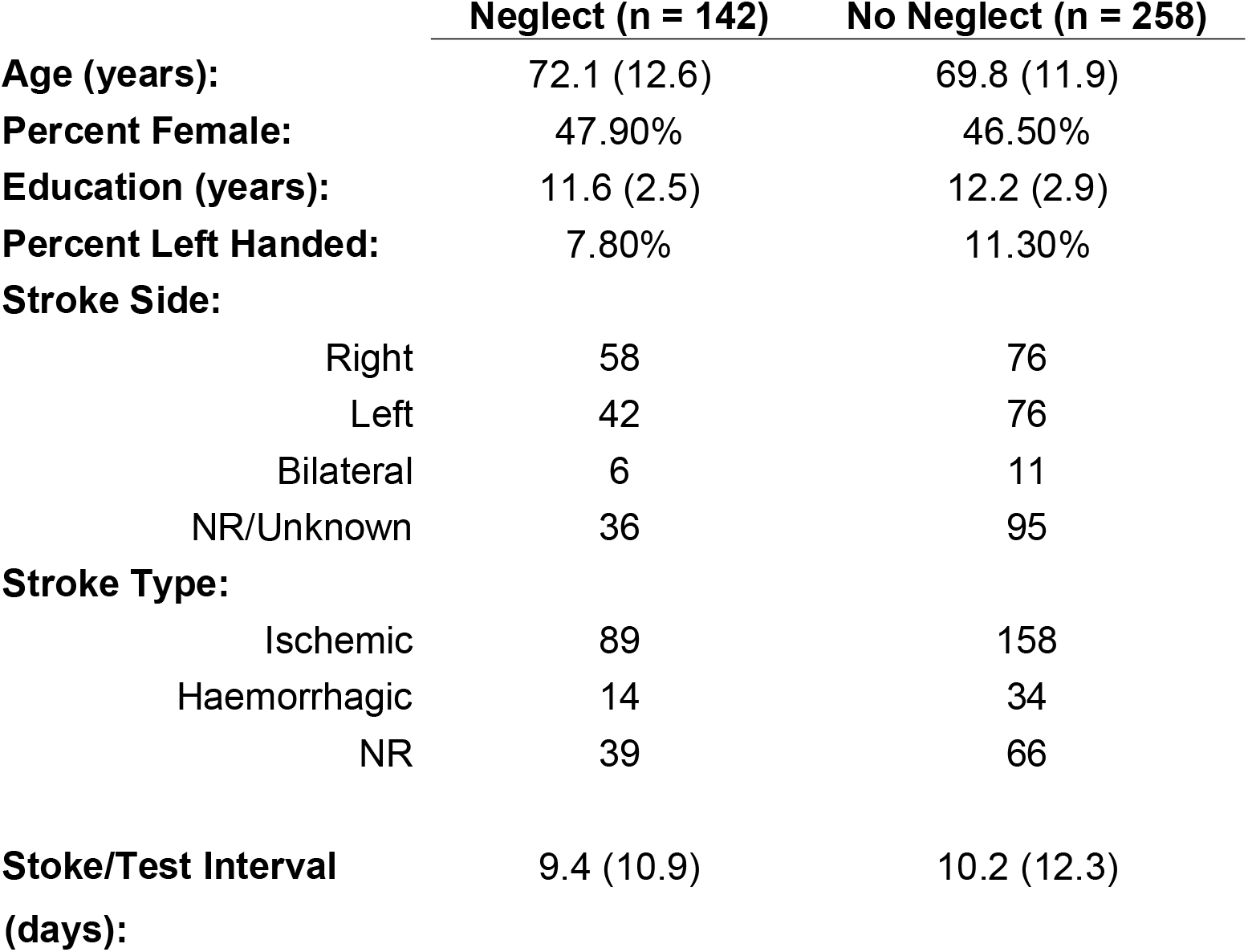
Demographics and Stroke Information for the patients with and without neglect. Means are reported alongside standard deviations (in parentheses).

Due to the high exclusion rate within this secondary analysis (60.8%), a series of analyses were conducted to determine whether the included neglect sample was representative of the full acute cohort assessed. The proportion of patients with neglect within all acute patients (n =1020, 44.5%) was found to be significantly higher than that within the included sample (n = 400, 35.5%) (X^2^= 5.579, p = 0.0182). However, there was no significant difference between the egocentric (t(265.79) = 0.627, p = 0.5312, 95% CI = - 0.974 – 1.884) or allocentric (t(230.24) = 0.720, p = 0.472, 95% CI = −0.563 – 1.211) asymmetry scores between neglect patients who were and were not included in the final sample.

### Procedures

The behavioural analyses conducted in this investigation aim to 1) determine which neglect factors predict presence of chronic neglect and 2) investigate the interaction between acute neglect type, lateralisation, severity with long-term SIS score. Data from standardised Cancellation Tasks and the Stroke Impact Scale questionnaire were considered in this investigation. Participants completed either the OCS or Birmingham Cognitive Screen Cancellation Task ^3,51^ in pen and paper format. These assessments are parallel versions of a standardised assessment aiming to detect egocentric and allocentric visuospatial neglect and do not aim to quantify additional neglect impairments (e.g. motor or auditory neglect). Each cancellation task consists of similarly structured search matrices containing complete, right-gap, and left-gap line drawings. In these tasks, patients were instructed to mark complete drawings while ignoring incomplete stimuli. Patients completed two practice trials and were given three minutes to complete the Cancellation Task. These Cancellation Tasks have been demonstrated to be sensitive and reliable measures for detecting and differentiating between allocentric and egocentric deficits ^3,36^ (Figure 2). The Stroke Impact Scale (SIS) is a self-report questionnaire assessing stroke-specific functional outcome domains^52,53^. This investigation employed version 3.0 of this scale, which includes 59 questions across 8 domains (physical weakness, memory, mood, communication, activities of daily living, mobility, affected hand, and participation). Each question is responded to on a 5-level Likert scale. The SIS exhibits high test-retest validity, correlates strongly with other established measures, and is sensitive to changes^35^.

**Figure 2:**
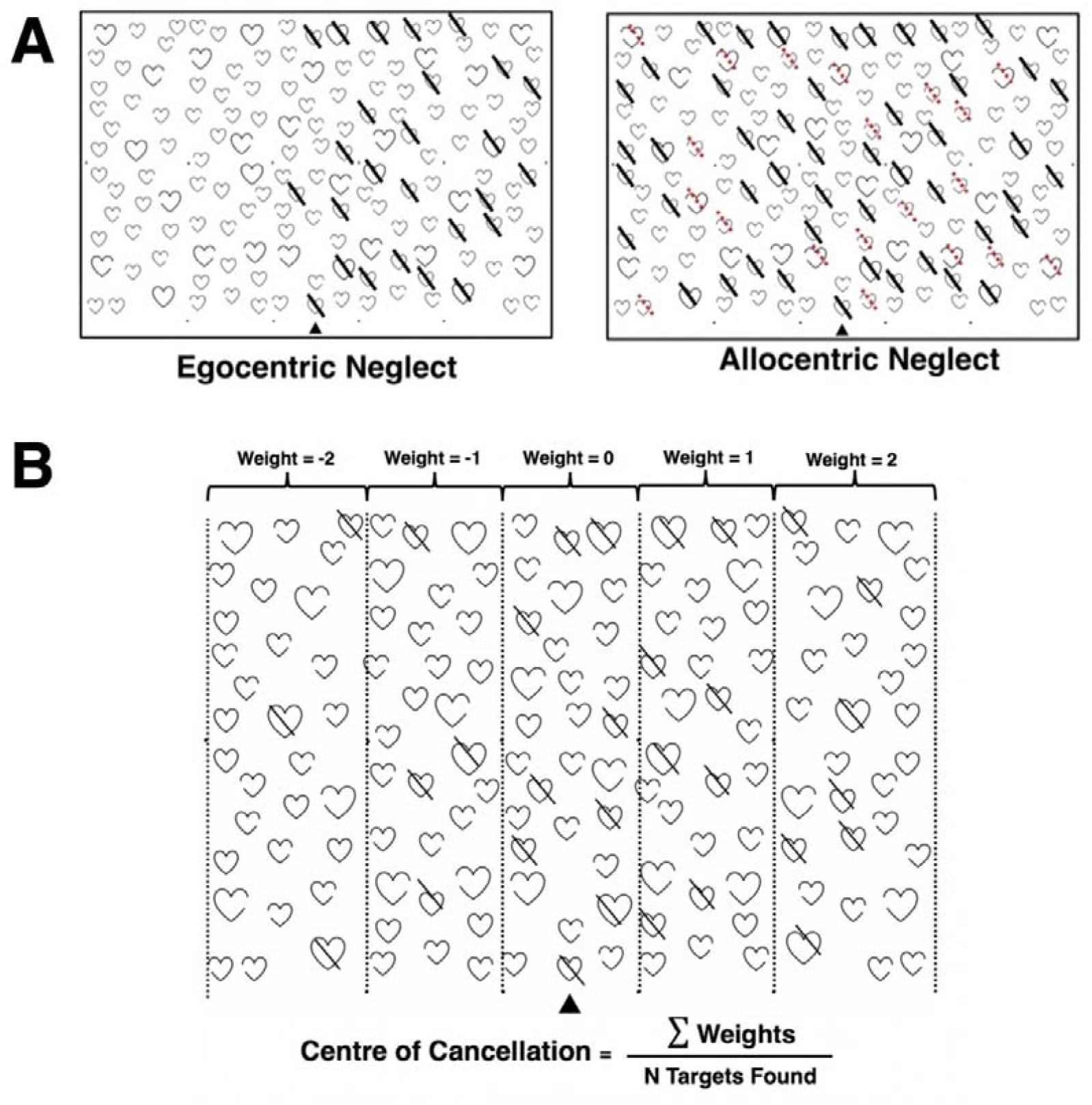
Neglect Impairment Definitions. A visualisation of egocentric and allocentric neglect deficits as detected by the OCS cancellation task (Panel A). Patients with egocentric neglect fail to report targets on one side of space while patients with allocentric neglect commit consistently lateralised false positive errors (highlighted in red). Panel B presents the task scoring grid, corresponding error weights, and equation for calculating centre of cancellation neglect severity scores.

Participants completed the Cancellation Task at timepoints 1 and 2. The presence of egocentric neglect is scored by subtracting the number of targets identified on the left from those on the right. Egocentric asymmetry more extreme than +3(left neglect)/-3(right neglect) in conjunction with total scores of less than 42/50 was considered to represent significant neglect impairment^36^. Egocentric neglect severity was calculated though a centre of cancellation (CoC) score^38-40^. This score was calculated by assigning each response a numerical weight according to its horizontal location^40^, the average response weight determines the CoC. Allocentric neglect impairment was quantified by subtracting the number of right-gap and left-gap false positives. Allocentric asymmetry more extreme than −1 (right neglect) or 1 (left neglect) represents significant impairment^49^. Allocentric severity is scored as the proportion of allocentric errors committed to number of targets successfully identified.

Participants also completed the SIS at timepoint 2. Total SIS score was calculated by formatting all Likert responses so that low scores represent poor outcomes (e.g. 1 = “could not do at all” and 5 = “not difficult at all”) and subsequently summing all numeric responses^52^. All data collected in this investigation has been made openly available on the Open Science Framework^54^ (https://osf.io/wm8v3/).

### Statistical Analysis

All comparisons control for age, sex, stroke-test interval, education, and stroke side when applicable. Given that patients with more severe strokes experience longer delays between admission and cognitive assessment, stroke-test interval was included as a proxy estimate of stroke severity.

First, two chi-squared tests were performed to determine whether the proportion of patients whose neglect impairment was unrecovered versus recovered at follow-up assessment was equivalent across neglect subtypes and lateralisations. Next, two t-tests (separately for egocentric and allocentric) were conducted to determine whether unrecovered patients initially had more severe impairments at T1. Finally, two repeated-measures ANOVAS (one for egocentric and allocentric) were performed to investigate whether the magnitude of the change in neglect severity between T1 and T2 is proportional for patients with different impairment lateralisation, and with/without chronic neglect. Each ANOVA included: presence/absence of chronic neglect, neglect lateralisation, age, sex, education, and stroke-test interval..

To investigate the second research question, a multiple regression was performed to determine the relationship between acute neglect severity and follow-up functional outcome (SIS score). Finally, an ANOVA was conducted to determine whether patients exhibiting different acute neglect subtypes/lateralisations reported significantly different follow-up SIS score.

## Results

### Neglect Prevalence and Recovery

142/400 (35.5%) patients exhibited significant neglect at T1. 71 (50%) of these cases involved only egocentric neglect, 41 (28.9%) only allocentric, and 30 (21.1%) cases involved co-occurring egocentric and allocentric impairments. 78 (54.9%) cases were left-lateralised, 57 (40.1%) impacted the right side of space, and 7(4.9%) involved right and left deficits which co-occurred^18,55^. There was no significant difference in stroke severity between patients with egocentric (mean stroke-test interval = 9.66 days) and allocentric (mean interval = 10.77 days) in this sample (t(83.50) = −0.474, p = 0.636, CI: −5.796 – 3.565). At follow-up, 98/142 (69%) neglect cases had fully recovered. There was no significant difference in the proportion of patients recovered between egocentric and allocentric neglect (X^2^(1)=0.66, *p=0.418*). Similarly, there was no significant association between the lateralisation of neglect and probability of neglect recovery (X^2^(2)=0.781, *p= 0.677)* (Figure 3).

**Figure 3:**
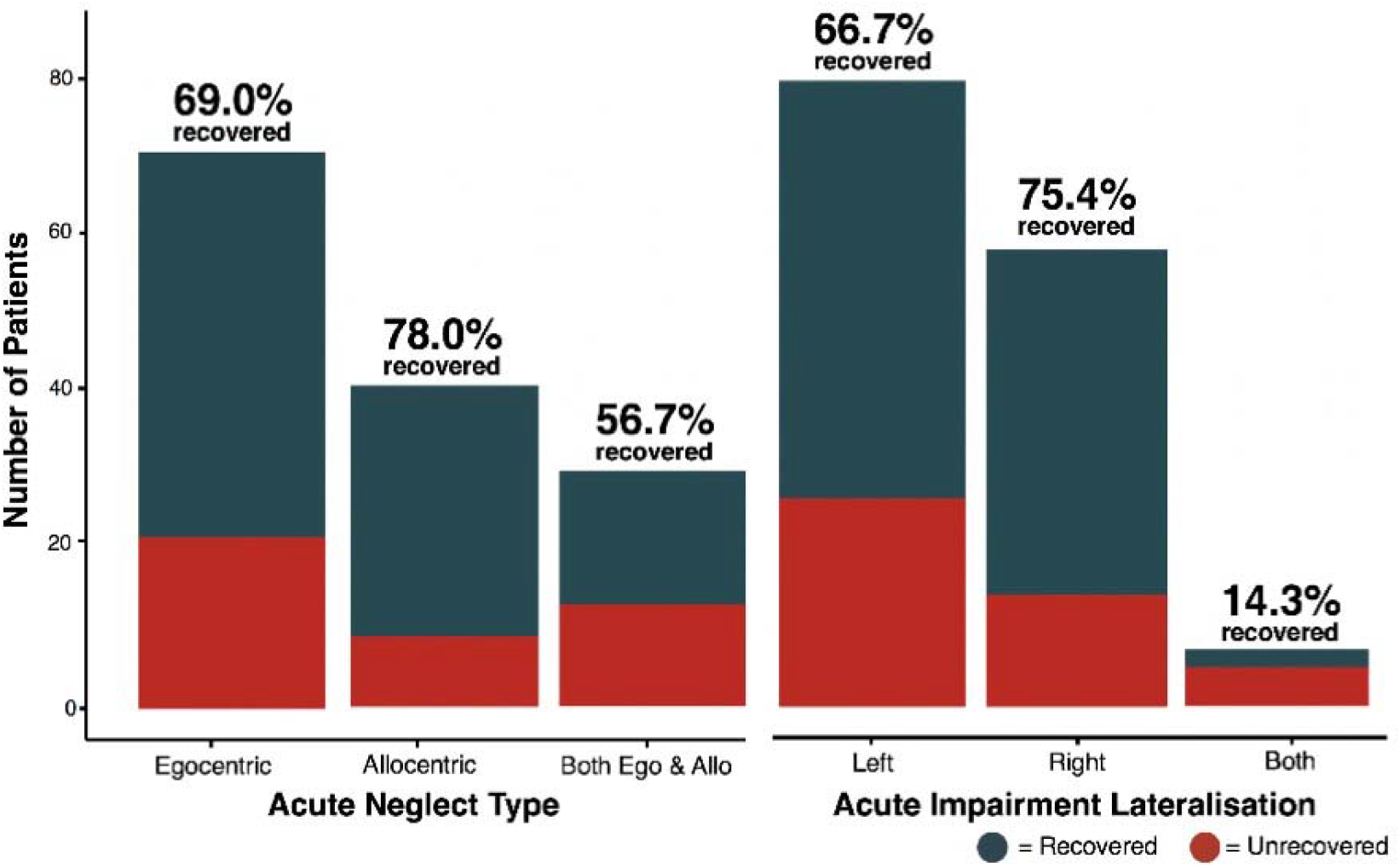
Neglect Subtype Recovery Proportions. Recovery proportions for each neglect subtype and for each lateralisation of neglect, demonstrating no reliable sub-group differences in proportion of neglect impairment at 6 months. Red areas represent patients who had not recovered by follow-up assessment.

Patients with chronic egocentric neglect had significantly more severe acute impairment than those who recovered, (t(34.933)= −2.288, p=0.028), suggesting that egocentric neglect follows a proportional recovery pattern. However, there was no difference in the severity of acute impairment between allocentric patients who did and did not recover (t(22.633= −1.373. p=0.183), suggesting that allocentric neglect follows a non-proportional recovery pattern.

Within egocentric neglect, there was a significant main effect of neglect lateralisation with left neglect being more severe than right neglect (initial CoC=0.721 vs. 0.348) (F(1,190)=13.912, p<0.001, *η*_*p*_^*2*^=0.068). However, no significant interaction effects were present between change in neglect severity over time and the presence of chronic neglect (F(1,190)=0.866, *p=0.353*) or between neglect lateralisation and the presence of chronic neglect (F(1,190)=2.218, p=0.138). These findings suggest that patients with egocentric neglect exhibit proportional recovery and that the rate of recovery is not different for patients with more and less severe neglect, regardless of lateralisation (see Figure 4).

**Figure 4:**
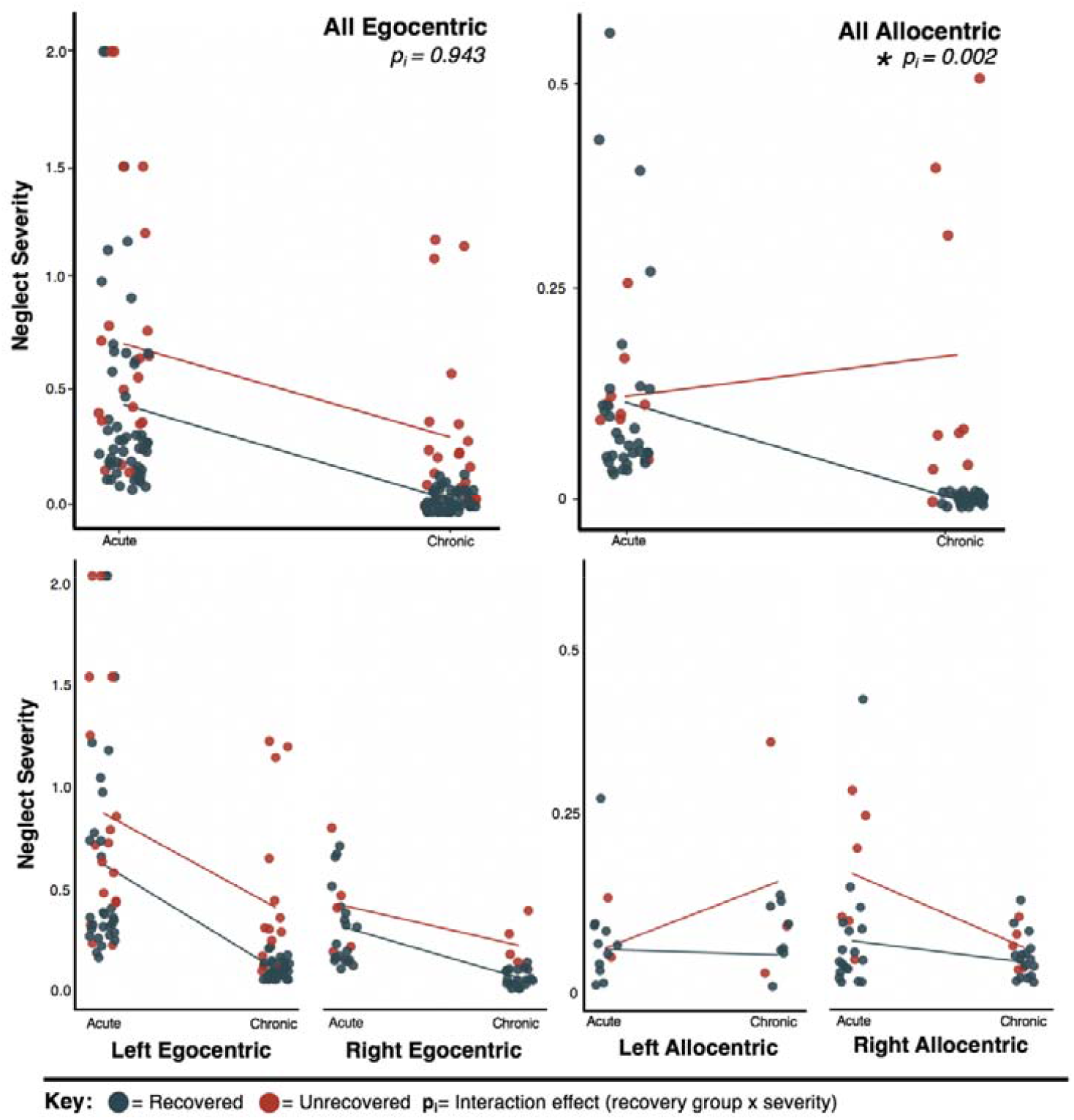
Proportional and Non-Proportional Recovery in Neglect. A visualisation of proportional recovery within patients with egocentric neglect and non-proportional recovery within patients with allocentric neglect. Dots represent patient neglect severity scores and lines visualise the difference between group means at each timepoint.

Within allocentric neglect, there was a significant main effect of impairment lateralisation on neglect severity (F(1,130)=7.759, p=0.006, η_p_^2^< 0.056), demonstrating that acute left allocentric neglect was significantly more severe than right allocentric neglect (severity=0.242 vs. 0.119). There was a significant interaction effect of change in neglect severity between timepoints and the presence of chronic neglect (F(1,130)=6.389, p= 0.017, η_p_^2^=0.047), demonstrating that allocentric cases do not recover at the same rate. Finally, there was no significant interaction effect between lateralisation and the incidence of chronic neglect (F(1,130)=2.218, p=0.138, η_p_^2^=0.001), suggesting that patients with right and left allocentric neglect do recover at similar rates.

### Which factors predict chronic SIS scores?

At the 6 month follow up, patients reported an average SIS total score of 223/300 (SD=44.3,Range=41-295), with lower scores indicating poorer outcome. A multiple regression was performed to investigate the relationship between neglect severity and functional outcome (SIS). This regression included egocentric and allocentric severity, impairment lateralisation, age, sex, test interval, handedness, and education as covariates (max VIF=1.27, Durbin-Watson=2.01). Overall, this model was found to be significant (F(9,383)=3.96, *p<0.001*, adjusted R^2^=0.066, Figure 5). Acute allocentric neglect severity (*p=0.031,Cohen’s f=0.109*), female sex (*p=0.003,Cohen’s f=0.154*), and test interval (*p=0.017,Cohen’s f=0.134*) were found to be significant predictors of lower functional outcome. Notably, acute egocentric severity did not act as a significant predictor of long-term SIS (p=0.083).

**Figure 5:**
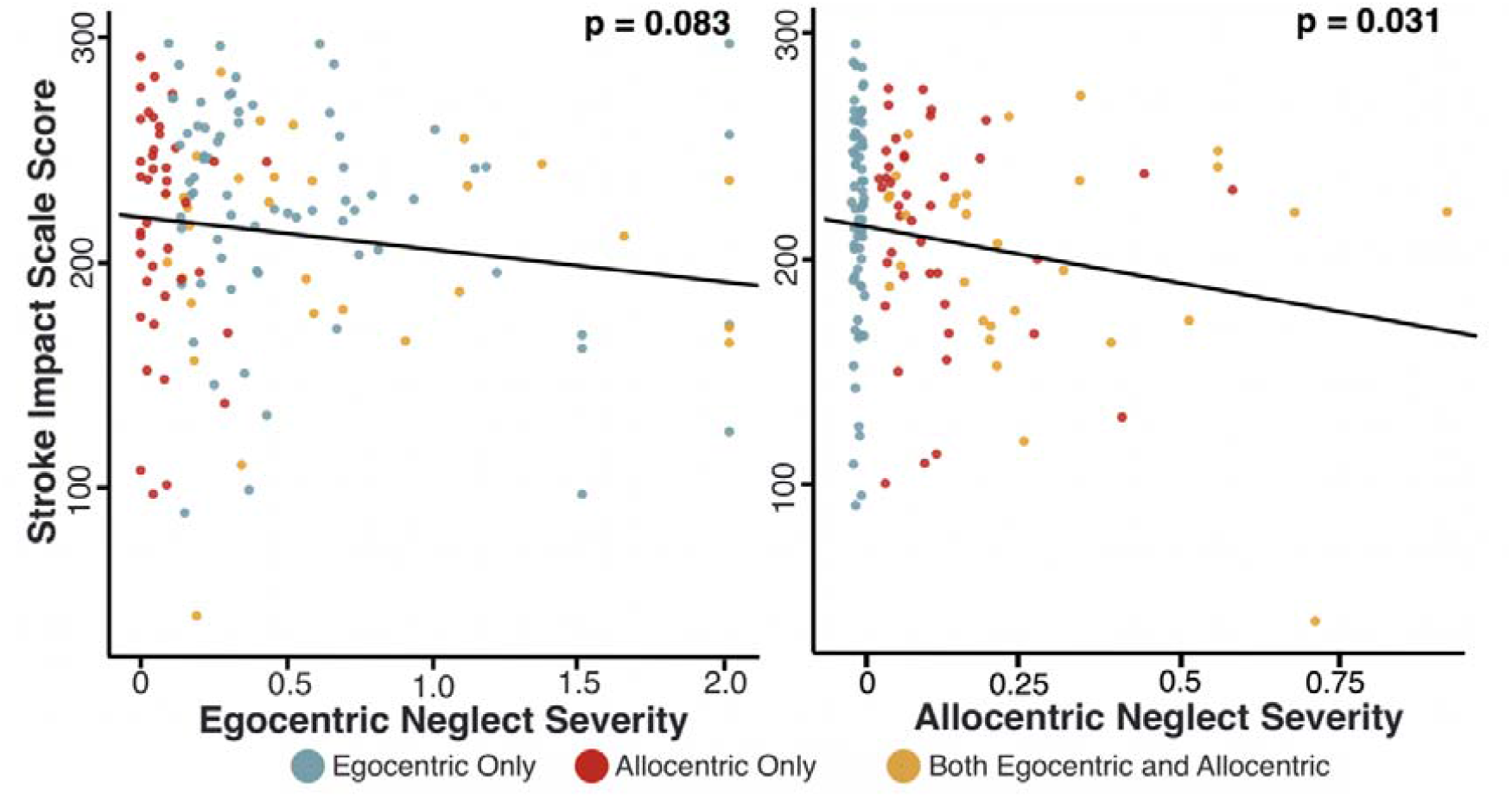
Neglect Severity and Functional Outcome. The relationship between egocentric neglect severity and allocentric neglect severity (as expressed through absolute values for centre of cancellation) and chronic SIS score. Patients with no neglect have not been plotted in this visualisation. See Figure 5 for an illustration of the distribution of SIS scores in patients with no neglect.

Finally, an ANOVA was conducted to investigate the interactions between acute neglect category/lateralisation and chronic SIS. There was a significant overall effect between neglect subtype and SIS score (F(9,383)=3.393, *p=0.018,η*_*p*_^*2*^=*0.026*). However, there were no significant post-hoc paired differences between different neglect categories (Figure 6).

**Figure 6:**
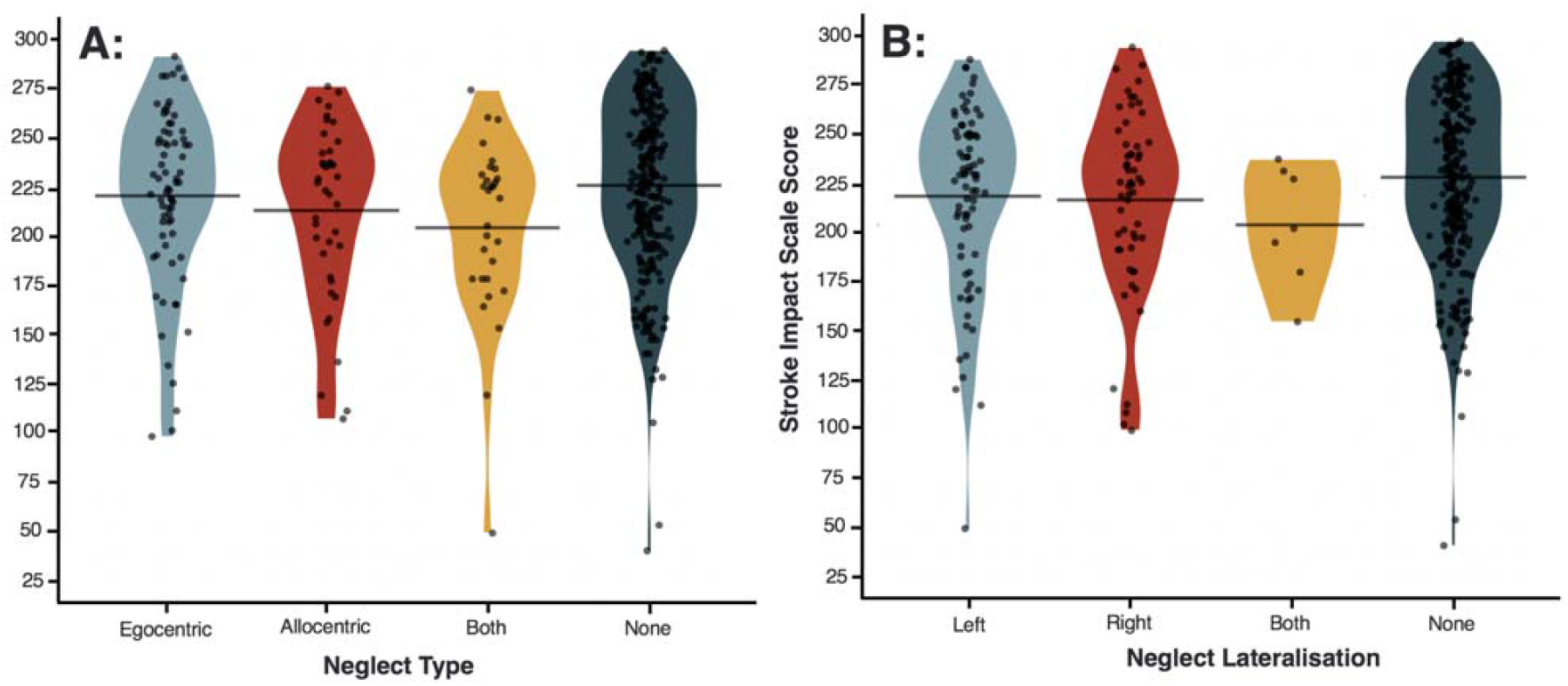
Functional Recovery by Neglect Subtype. The interaction between acute neglect category (Panel A) and lateralisation (Panel B) and chronic SIS score.

## Discussion

This study aimed to determine whether visuospatial neglect subtypes act as predictors of long-term outcomes, both with regards to neglect recovery and to broad functional outcomes. Egocentric and allocentric neglect occurred frequently in the acute stroke population and represented doubly dissociated conditions. Both left- and right lateralised neglect deficits were common and, in line with expectations^40^, left-lateralised neglect was more severe than right neglect. Egocentric neglect exhibited proportional recovery with the severity of acute impairment predicting the severity of chronic impairment. However, allocentric neglect was not found to recover proportionally with a subset of patients remaining unchanged or getting worse over time, regardless of initial severity. The severity of acute allocentric, but not egocentric, neglect acted as a significant predictor of long-term functional outcome. These results highlight the importance of standardised, neuropsychological assessment of post-stroke visuospatial impairments to improve the sensitivity of neglect detection and provide quantitative severity information which can be used to predict functional outcome trajectory and guide intervention pathways.

The results of the present study suggest that egocentric neglect’s initial severity modulates the probability of its recovery. Egocentric patients who recovered had less severe acute neglect than patients presenting with chronic neglect at 6 months. Importantly, the rate of egocentric improvement was not different between patients whose impairments did and did not recover. This effect held for patients exhibiting right and left egocentric neglect.

These findings align with previous studies documenting proportional recovery in egocentric neglect and suggesting that acute severity may be the best indicator of the probability of egocentric neglect recovery^39,45^.

However, proportional recovery was not found to be present within patients with allocentric neglect. Allocentric patients who recovered by 6 months were not initially less severely impaired than patients whose allocentric neglect did not recover. This pattern is characteristic of non-proportional recovery^46–48^. This non-proportional recovery pattern within allocentric neglect aligns well with recent research suggesting that not all post-stroke impairments follow the proportional recovery rule^47,48^. Additionally, unrecovered allocentric patients were found to exhibit a significantly different recovery rate than patients whose allocentric neglect did recover. Considered cumulatively, these findings suggest that it is not yet clear how to predict which patients with allocentric neglect will and will not recovery from their neglect impairment. Future research can aim to investigate the recovery of allocentric neglect in more detail, as it appears that this subtype of neglect is more likely to lead to a chronic neglect impairment. Similarly, it is important for future studies to aim to identify additional factors such as behavioural profile or comorbid cognitive impairments which may help distinguish between allocentric neglect patients who do and do not spontaneously recovery.

This investigation also considered the impact of neglect lateralisation and severity on functional outcome. With regards to lateralisation, interestingly, there was no significant difference in functional impairment levels reported by patients with right and left lateralised neglect, despite the finding that left neglect was significantly more severe than right neglect on neuropsychological assessments. This effect has been previously identified^40^, but it is not clear which factors are responsible for driving it. It seems plausible that right neglect may result in a disproportionally high impact on functional activity relative to impairment severity, as left hemisphere damage may be more likely to reduce patient’s ability to use their dominant hand, and include language and communication impairments which may affect overall functional outcomes in a broad measure such as the SIS. Further research is needed to investigate this possibility in detail.

The quantitative severity of acute allocentric neglect impairment was found to act as a significant predictor of poor long-term functional outcome while the severity of acute egocentric neglect was not. Patients with more severe allocentric neglect at the acute timepoint were found to report higher levels of domain-general functional impairment at chronic assessment, regardless of whether or not their neglect had recovered. This effect could potentially be explained by a comparatively more severe impact of allocentric neglect impairment on daily life activities. Patients with egocentric neglect may be able to orient or adjust their environments to help compensate for their neglect, but it is unclear whether patients with allocentric-level deficits would benefit from similar compensation strategies. Similarly, allocentric neglect may impact performance within specific activities (e.g. reading individual words, identifying/manipulating objects) which may be comparatively unaffected by egocentric neglect. Additional research is needed to clarify how egocentric and allocentric neglect deficits differentially impact on functional activities.

Considered cumulatively, the findings of this study emphasize the importance of employing standardised, neuropsychological assessments of neglect within clinical environments. Detecting and determining the severity of allocentric neglect can help identify patients which are least likely to spontaneously recover and are most likely to experience lower levels of functional outcome, but this impairment is not commonly screened for after stroke. However, many commonly used neglect assessments are not able to reliably detect allocentric neglect, resulting in the omission of information which can help identify patients likely to experience poorer functional outcomes. Implementing standardised administration of neuropsychological neglect assessments is critically important in order to sensitively detect neglect impairments, to assign quantitative neglect severity scores, and to identify neglect patients who are most likely to benefit from targeted neglect-specific rehabilitation programmes ^3,16,49^. Based on the current data, neglect intervention research may need to focus on rehabilitation specifically for those with more severe neglect impairments and those with allocentric neglect.

This investigation also highlights the need for future neglect research to accurately represent the diversity of neglect behavioural impairments to produce generalisable findings. Studies which consider only a restricted subset of neglect impairment (e.g. restricting sample to only left egocentric neglect after right hemisphere damage) are likely not representative of neglect as a whole and as it presents in acute clinical settings. Differences in patient inclusion criteria can help explain why many past investigations have provided conflicting findings pertaining to the prevalence ^5,56,57^, underlying neural mechanisms ^12,58,59^, and functional impact of neglect impairment ^6,7,10^. Similarly, future investigations should aim to determine whether neglect uniformly impacts across all aspects of functional recovery, or whether a single component of this broad, encompassing concept (e.g. activities of daily living, mood) may be responsible for driving the predictive relationship between neglect and poor functional outcome. It is therefore critically important for future neglect research to adequately represent this syndrome’s behavioural diversity as conceptualising neglect as a unitary syndrome rather than a cluster of interrelated impairments may preclude valid conclusions about the disorder as a whole.

Most importantly, the findings of this investigation reveal that patients with allocentric neglect may represent a key group for research for targeted, neglect-specific rehabilitation. The vast majority of existing neglect rehabilitation strategies are designed to ameliorate the effects of egocentric neglect ^60–62^. It seems likely that many of these strategies would be ineffective for patients with allocentric impairment. For example, rehabilitation strategies such as prism adaptation help shift attention towards a neglected hemifield but would likely not impact an attentional bias which is not mediated by viewer-centred impairments ^60^. Pharmaceutical and transcranial stimulation based rehabilitation strategies may face similar challenges ^62^. However, some rehabilitation studies which could plausible impact the severity of both egocentric and allocentric neglect have been proposed^62^.

Approaches based on phasic alerting^63^, vestibular stimulation^64^, neck muscle vibration^65^, or motor feedback training^66^ seem potentially appropriate for ameliorating allocentric neglect symptoms. However, the impact of these rehabilitation therapies on allocentric-level attentional biases have not yet been adequately investigated in large and representative samples of neglect patients. It is not clear how allocentric neglect impacts behaviour, and understanding this impact is likely a necessary precursor to identifying effective rehabilitation strategies.

### Limitations

SIS scores are self-reported and are therefore vulnerable to response biases. In addition, some visuospatial neglect patients may also exhibit anosognosia^67^. Nevertheless, this association may influence functional self-report data and lead to some underestimation of functional impairment levels. Given that this study is a secondary analysis, the large sample data came at the cost of detailed stroke and additional potentially informative data. For example, T1 functional assessments, tests for additional neglect signs (e.g. anosognosia), rehabilitation programme information, detailed stroke anatomy, or diagnoses of additional neglect subtypes were not available.

Importantly, egocentric and allocentric neglect are not the only subtypes of visuospatial neglect. Given that this investigation demonstrated that egocentric and allocentric neglect may follow different recovery trajectories, it seems plausible that other neglect subtypes, not considered within this investigation, may be differentially associated with long term recovery as well. Future research should therefore aim to elucidate the recovery trajectories associated with additional visuospatial neglect subtypes. Critically, this investigation aimed to determine predictors rather than causes of poor recovery outcome. The identified significant relationship between acute allocentric neglect and poor recovery outcome should therefore not be interpreted as a causal relationship but instead as a correlational effect which may help clinicians predict which patients may experience reduced functional outcome in later stroke recovery phases. A number of factors including overall stroke severity, age of onset, co-morbid impairments, cognitive reserve, and psychosocial factors have been shown to be significantly related to stroke recovery trajectories^68–70^. For this reason, any individual factor, including allocentric neglect severity, should not be understood as the single predictor of stroke recovery but instead only explains a small portion of variance (approximately 7%). Clinicians should therefore employ a wholistic approach considering the impact multiple factors such as overall stroke severity, age of onset, comorbid impairments, cognitive reserve, and psychosocial factors rather than any one source of information to make informed prognoses. Importantly, this study does not aim to develop a quantitative prognostic model, but instead to qualitatively identify groups of patients who are less likely to spontaneously recover over time. Future research is needed to develop and validate more precise, quantitative prognostic models

Finally, the results of any individual analysis are determined by the specific combination of inclusion criteria, impairment definitions, and outcome measures employed^71,72^. Additional research employing multiverse analysis techniques is needed to determine the degree to which the findings of this investigation are robust and generalisable across many different potential patient groups and outcome measures (e.g. SIS sub-domains) (see Moore & Demeyere (Under Review)^73^).

### Conclusion

The findings of this investigation illustrate that considering the subtype and quantitative severity of visuospatial neglect provides information which can be applied to identify patients who are more likely to end up living with chronic neglect. These findings highlight the need for standardised neuropsychological neglect assessments and for the development of allocentric-specific rehabilitation strategies.

## Data Availability

All data collected in this investigation has been made openly available on the Open Science Framework (https://osf.io/wm8v3/).

## Acknowledgements

This work was funded by Stroke Association UK awards to ND (TSA2015_LECT02; TSA 2011/02), MJM (SA PGF 18\100031), KV (TSA PDF 2017/03), the Wellcome Trust award to CRG (101253/A/13/Z) and was supported by the National Institute for Health Research (NIHR) Oxford Biomedical Research Centre (BRC) based at Oxford University Hospitals NHS Trust. We would like to express our sincere gratitude and admiration to the late Prof Glyn W Humphreys, who initiated the OCS work and led the OCS-Care study. The OCS study was supported by the National Institute for Health Research Clinical Research Network. We also acknowledge the contributions to data collection and curation for the OCS data made by Ms Ellie Slavkova, Ms Grace Chiu and Ms Romina Basting.

